# Monoallelic *de novo* variants in *DDX17* cause a novel neurodevelopmental disorder

**DOI:** 10.1101/2023.10.11.23295963

**Authors:** Eleanor G. Seaby, Annie Godwin, Valentine Clerc, Géraldine Meyer-Dilhet, Xavier Grand, Tia Fletcher, Laloe Monteiro, Valerio Carelli, Flavia Palombo, Marco Seri, Giulia Olivucci, Mina Grippa, Claudia Ciaccio, Stefano D’Arrigo, Maria Iascone, Marion Bermudez, Jan Fischer, Nataliya Di Donato, Sophie Goesswein, Marco L. Leung, Daniel C. Koboldt, Cortlandt Myers, Dennis Bartholomew, Gudny Anna Arnadottir, Kari Stefansson, Patrick Sulem, Ethan M. Goldberg, Ange-Line Bruel, Frederic Tran Mau Them, Marjolaine Willems, Hans Tomas Bjornsson, Hakon Bjorn Hognason, Eirny Tholl Thorolfsdottir, Emanuele Agolini, Antonio Novelli, Giuseppe Zampino, Roberta Onesimo, Katherine Lachlan, Diana Baralle, Heidi L. Rehm, Anne O’Donnell-Luria, Julien Courchet, Matt Guille, Cyril F. Bourgeois, Sarah Ennis

## Abstract

**Introduction:** DDX17 is an RNA helicase shown to be involved in critical processes during the early phases of neuronal differentiation. Globally, we identified 11 patients with neurodevelopmental phenotypes with *de novo* monoallelic variants in *DDX17*. All 11 patients had a neurodevelopmental phenotype, whereby intellectual disability, delayed speech and language, and motor delay predominated.

**Materials and methods:** We performed *in utero* cortical electroporation in the brain of developing mice, assessing axon complexity and outgrowth of electroporated neurons, comparing wild-type and Ddx17 knockdown. We then undertook *ex vivo* cortical electroporation on neuronal progenitors to quantitively assess axonal development at a single cell resolution. Homozygous and heterozygous *ddx17* crispant knockouts in *Xenopus tropicalis* were generated for assessment of morphology, performed behavioural assays, and neuronal outgrowth measurements. We further undertook transcriptomic analysis of neuroblastoma SH-SY5Y cells, to identify differentially expressed genes in DDX17-KD cells compared to controls.

**Results:** Knockdown of Ddx17 in electroporated mouse neurons *in vivo* showed delayed neuronal migration as well as decreased cortical axon complexity. Mouse primary cortical neurons revealed reduced axon outgrowth upon knockdown of *Ddx17 in vitro*. The axon outgrowth phenotype was replicated in crispant *ddx17* tadpoles, including in a heterozygous model. Crispant tadpoles had clear functional neural defects and showed an impaired neurobehavioral phenotype. Transcriptomic analysis identified a statistically significant number of differentially expressed genes involved in neurodevelopmental processes in DDX17-KD cells compared to control cells.

**Discussion:** We have identified a new gene, *DDX17*, representing a rare cause of neurodevelopmental delay. We provide evidence for the role of the gene and mechanistic basis of dysfunctional neurodevelopment in both mammalian and non-mammalian species.

## Introduction

RNA helicases have essential biochemical roles in all aspects of RNA metabolism, including unwinding and annealing RNA molecules and remodelling ribonucleoprotein complexes. The DEAD-box proteins are highly conserved across species, comprising the largest family of RNA helicases.^1,2^ They share twelve conserved motifs including the signature DEAD motif compromising the amino acid sequence Asp-Glu-Ala-Asp. DDX17, also known as DEAD box protein 17, (and its close homolog DDX5), are highly energy dependent DEAD-box RNA helicases involved in diverse cellular processes, notably gene expression, biogenesis of miRNAs via their interaction with the Drosha/DGCR8 complex, and the regulation of cell fate switches and biological transitions.^3,4^ They are coregulators of several transcription factors including MYOD, a master regulator of muscle differentiation and SMAD proteins, which mediate transforming growth factor beta induced epithelial-to-mesenchymal transition.^4,5^ Additionally, they are components of the spliceosome and regulate alternative splicing.

*DDX17*, located on chromosome 22q13.2, has been shown to be involved in the control of Repressor Element 1-silencing transcription factor (REST) related processes that are critical during the early phases of neuronal differentiation.^6^ Through its association with REST, DDX17 promotes its binding to the promoter of certain REST-targeted genes and coregulates the transcriptional repression activity of REST. DDX17 and the REST complex are downregulated during neuroblastoma cell differentiation, affecting activation of neuronal genes. Furthermore, DDX17 and DDX5 regulate the expression of multiple proneural microRNAs which target the REST complex during neurogenesis, implicating DDX17 in neuronal gene repression.^6^ In 2022, Suthapot *et al.*^7^ focused on characterising chromatin occupancy of DDX17 and DDX5 in hPSCs NTERA2 and their neuronal derivates. They showed that the expression of both helicases is abundant throughout neural differentiation of the hPSCs NTERA2, preferentially localised within the nucleus and that they occupy chromatin genome-wide at regions associated with genes related to neurogenesis. Both DDX17 and DDX5 are mutually required for controlling transcriptional expression of these neurogenesis-associated genes but are not important for maintenance of the stem cell state of hPSCs. In contrast, they are critical for early neural differentiation of hPSCs, possibly due to their role in the upregulation of key neurogenic transcription factors such as SOX1, SOX21, SOX2, ASCL1, NEUROG2 and PAX6. Critically, DDX17 and DDX5 are important for differentiation of hPSCs towards NESTIN and TUBB3 positive cells, which represent neural progenitors and mature neurons, respectively. However, those studies used a DDX17 and DDX5 co-depletion approach to address the function of these factors in neurogenesis, and information regarding the specific contribution of each helicase to this process is lacking.

To date, *DDX17* has no disease-gene relationship. The gene is highly constrained for loss-of-function (LoF), i.e. fewer LoF variants in *DDX17* are observed in population datasets than would be expected under a null mutational hypothesis (37.7 expected, 1 observed, pLI=1.0 in gnomAD). Genes can be quantified by constraint to LoF using the Loss-of-function Observed/Expected Upper-bound Fraction (LOEUF) score, which places genes along a continuous spectrum of intolerance to haploinsufficiency.^8^

Genes highly constrained for LoF, represented by low LOEUF scores, are highly associated with known haploinsufficient disease genes.^8,9^ However, the majority of genes in the lowest LOEUF decile are not yet associated with a disease phenotype but may be expected to cause disease if mutated through LoF.^10^ *DDX17* has a LOEUF score of 0.13, suggesting that haploinsufficiency of the gene is not tolerated.

Considering its role in neuronal differentiation, muscle differentiation and alternative splicing, one might expect that *DDX17* is an essential gene in neurodevelopment and may present such a phenotype in humans. Therefore, identification of patients with LoF variants in *DDX17* may help characterize a new gene-disease relationship.

## Methods

### Accessing patient data

We obtained access to the 100,000 Genomes Project data through membership of a Genomics England Clinical Interpretation Partnership, with approved project RR359: Translational genomics: Optimising novel gene discovery for 100,000 rare disease patients. Deidentified whole genome sequencing and phenotype data (stored as human phenotype ontology terms) were accessible in the Genomics England Research Environment.

Additional study subjects were identified through Matchmaker Exchange after deposition in GeneMatcher.^11,12^ In total, 11 patients consented to participate in our study. Parents and legal guardians of all affected individuals provided written consent for the publication of their results alongside genetic and clinical information. Guardians of patients 6, 7 and 10 explicitly consented to have photographs published.

### Sequencing and data analysis

All patients, except for patient 3, had trio exome sequencing performed. Data processing and variant filtering and prioritization were carried out by in house pipelines at respective host centres. Patient 3 had trio whole genome sequencing undertaken as part of the 100,000 Genomes Project^13^ and their data was filtered using the DeNovoLOEUF filtering strategy.^9^ DeNovoLOEUF is a tool that can be applied at scale to genomics datasets, extracting rare *de novo* predicted loss-of-function variants in LOEUF-constrained genes.^9^

### Xenopus tropicalis

Adult Nigerian strain *Xenopus tropicalis* were housed within the European *Xenopus* Resource Centre (E*X*RC; https://xenopusresource.org), University of Portsmouth, in recirculating MBK Ltd systems maintained at 24°C - 27°C (13-11-hour light-dark cycle) with 10% daily water changes. All *Xenopus* work was completed in accordance with the Animals (Scientific Procedures) Act 1986 under licence PP4353452 following ethical approval from the University of Portsmouth’s Animal Welfare and Ethical Review Body. Detailed methods on: the generation of *X. tropicalis* founder animals and ddx17 crispants; wholemount *in situ* hydridisation; phenotypic analysis; experimental design and statistical analysis are available in **Supplementary methods**.

### RNA-seq

Detailed methods are available in **Supplementary methods**. In summary, human SH-SY5Y neuroblastoma cells (ECACC #94030304) were grown and transfected with 60 nM of a mixture of 2 different siRNA against *DDX17* (Merck-Millipore, see sequences in **Table S1**). Protein extraction was carried out as previously described^14^ and total RNA were isolated. Directional RNA libraries were prepared from total RNA after removal of ribosomal RNA (lncRNA library, Novogene). High throughput sequencing of 150 bp paired-end reads was carried out on an Illumina Novaseq 6000 platform (Novogene), generating an average number of 75 million matched pairs of reads per sample. Raw reads were pre-processed and mapped reads were filtered using SAMtools^15^ and the number of reads per gene was counted using HTSeq.^16^ Differential gene analysis was carried out with the DESeq2^17^ package. Parameters for differential expression: P-value < 0.05, [log2(FC)] ≥ 0.50 and base mean ≥ 10. Gene ontology and gene-set enrichment analyses were carried out using the ShinyGO 0.76.3 web interface.^18^

### Mouse experiments

Mouse breeding and handling was performed according to experimental protocols approved by the CECCAPP Ethics committee (C2EA15) of the University of Lyon, and in accordance with French and European legislation. Detailed methods on *ex vivo* cortical electroporation and primary neuronal cultures; immunostaining; *in utero* cortical electroporation; immunohistochemistry; image acquisition; and quantifications and statistical analyses are available in **Supplementary methods.**

## Results

We applied the DeNovoLOEUF filtering strategy, as previously described^9^, to 13,494 parent/offspring trios in the 100,00 Genomes Project, focusing on genes with a LOEUF score <0.2 with no prior disease gene association. We identified one individual harboring a heterozygous pLoF variant in *DDX17*. Using the GeneMatcher platform, we identified a further 10 patients all with *de novo* variants in *DDX17* presenting with neurodevelopmental phenotypes. Representatives for these participants were then invited to join our research study and referring clinicians were asked to complete a standardized phenotype table (**Supplementary File B**). The summary of the phenotypic features of the 11 patients (from 11 independent families) harbouring *de novo* heterozygous variants in *DDX17* are provided in **Table 2**. All variants were absent from gnomAD^8^ v2.1.1 and v.3.1.2.

**TABLE 2.**
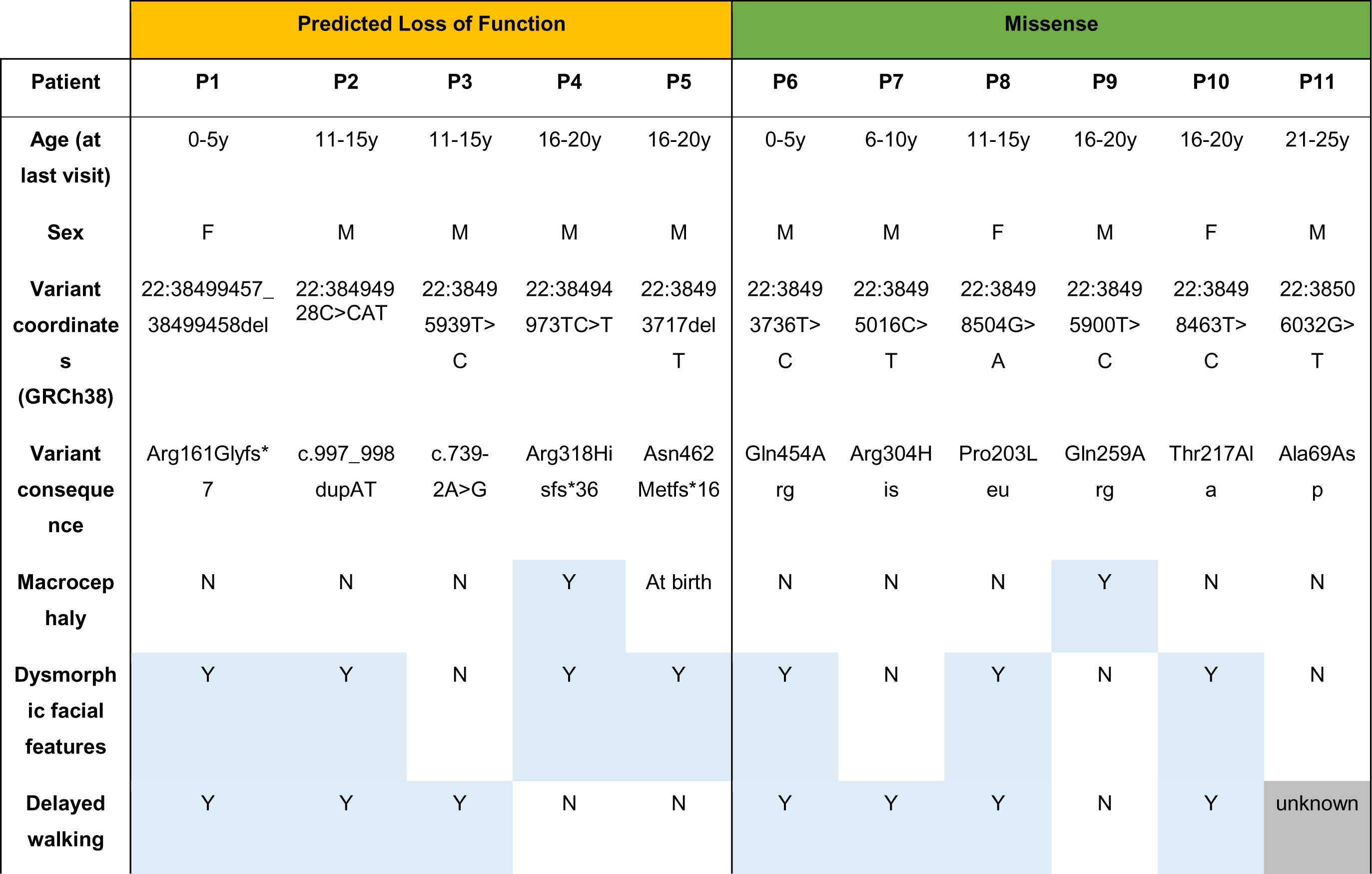
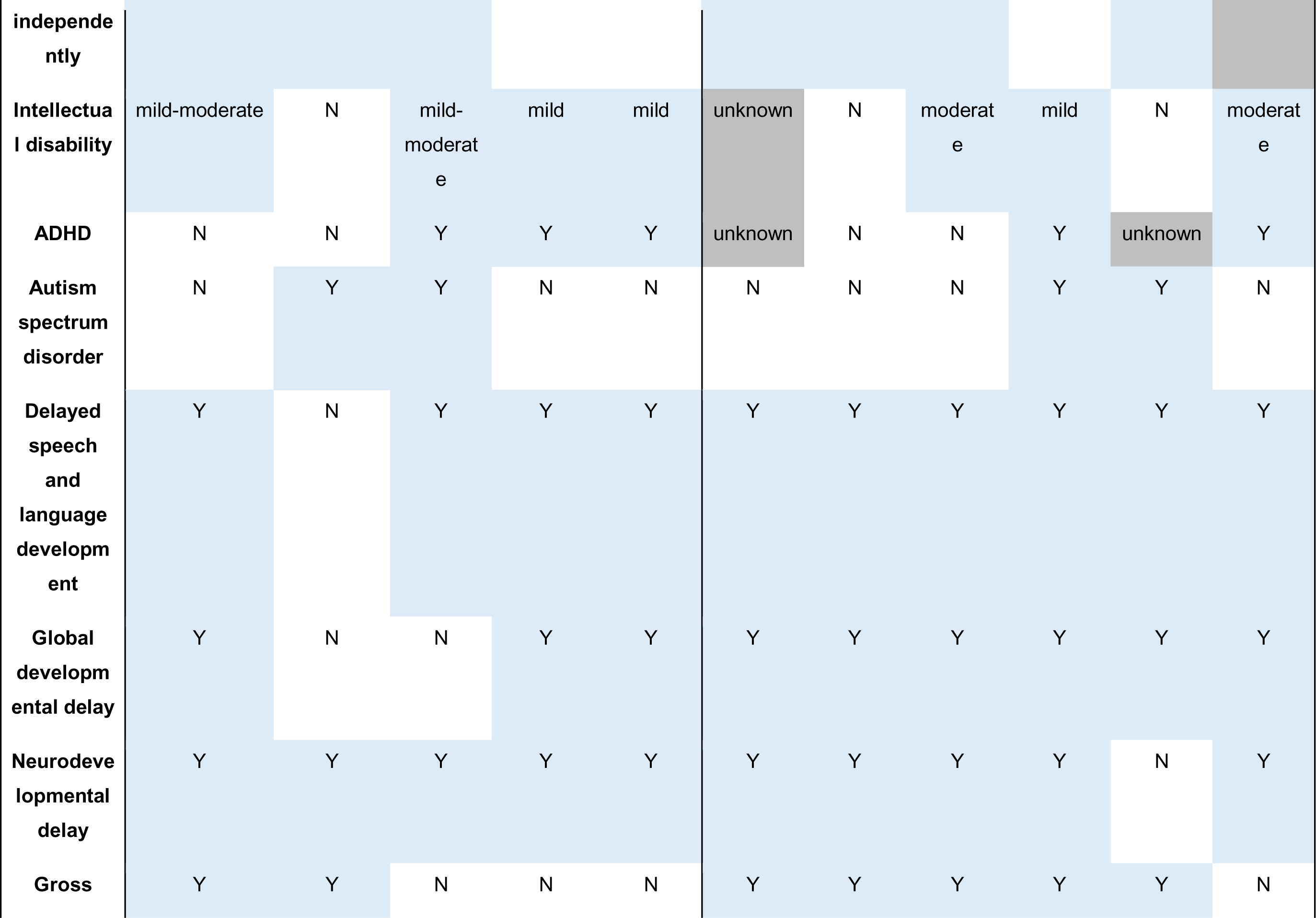
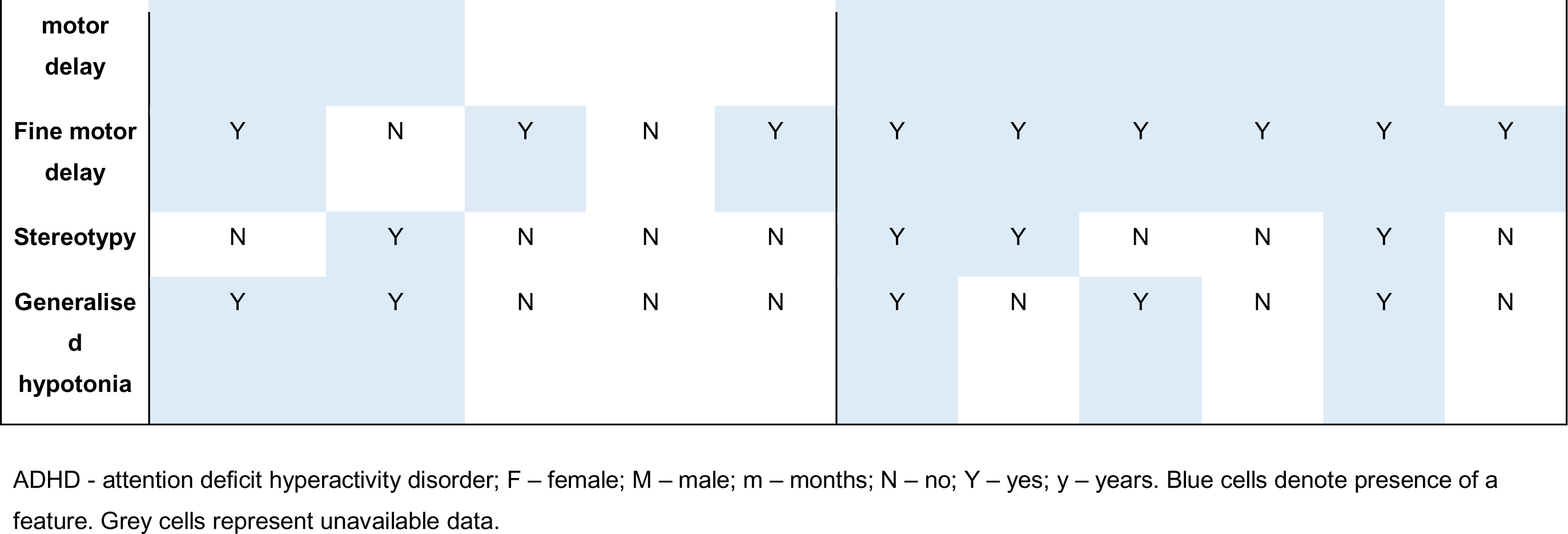
Core phenotypic features of cohort with heterozygous *de novo* variants in *DDX17*.

The cohort comprises 8 males and 3 females, all of whom are alive and have a median age of 15y at the latest available follow up. The median age of walking was 18 months. Intellectual disability, ranging from moderate to mild (IQ 56-83), is prevalent in 7/10 (70%) of patients. Seven of eleven (64%) of the cohort have dysmorphic facial features. Overlapping facial dysmorphology between patients includes: telecanthus; synophrys; upslanting palpebral fissures; depressed nasal bridge; posteriorly rotated ears; high arched eyebrows; epicanthus; telecanthus; frontal bossing; micrognathia and strabismus. Three patients have 5^th^ finger clinodactyly, two have clubfeet, and three have 2,3 toe syndactyly (some of these features are shown in **Figure 1A**). Fifty-six percent (5/9) have attention deficit hyperactivity disorder (ADHD) and 4/11 (36%) have features of autism. Ninety-one percent (10/11) have delayed speech and language development, 9/11 (82%) have global developmental delay, and 11/11 (100%) have neurodevelopmental delay. Gross motor delay is prevalent in 8/11 (73%), and 9/11 (82%) have fine motor delay. Thirty-six percent (4/11) have stereotypy and 5/11 (45%) have generalized hypotonia. Patient 9 (height 178cm (Z=0.40); weight, 55.7kg (Z= -0.89)) and patient 4 (height 165cm (7th percentile); weight, 51.5kg (4^th^ percentile)) have signs of macrocephaly with Z-scores of 3.97 and 3.23, respectively. Patient 5 had signs of macrocephaly at birth, which normalized through infancy. Eight participants had brain MRI scans of which 4 patients showed abnormalities including: left lateral compartment greater than right; monolateral temporal cortical dysplasia; asymmetry of the cerebral cortex and right sided nonspecific demyelination; generalized brain demyelination, and periventricular white matter hyperintensities. No other obvious asymmetry was observed.

**FIGURE 1.**
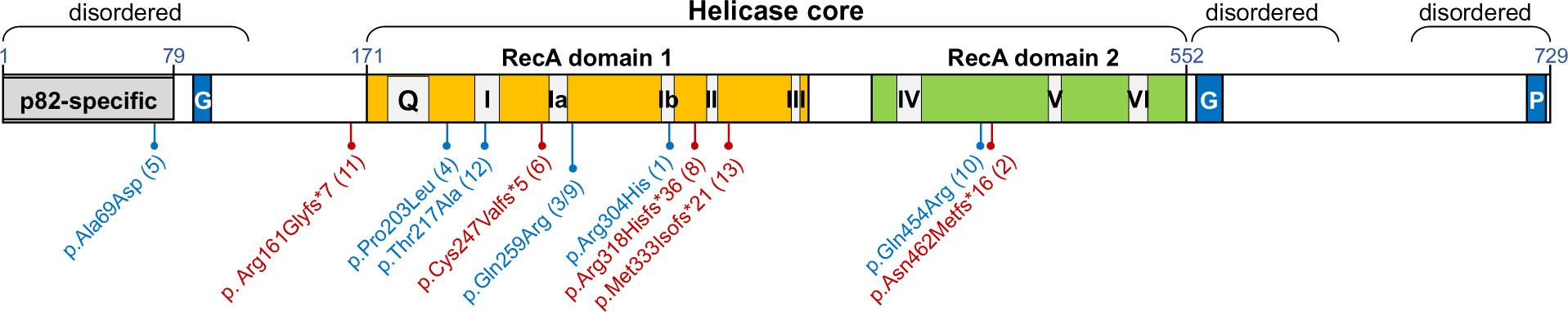
DDX17 patient variants. Gene ideogram, whereby variants in blue are missense, and variants in red are loss of function.

### Molecular genetic findings

Following variant filtering and prioritization of exome and genome data, no likely pathogenic or pathogenic variants as curated using American College of Medical Genetics and Genomics and the Association for Molecular Pathology (ACMG-AMP) guidelines^19^ were identified that fully explained the patients’ phenotypes. Eleven participating research programs identified *de novo* variants of uncertain significance in *DDX17* that were of sufficient interest to submit to Matchmaker Exchange. Five variants were pLoF and 6 variants were missense (**Figure 1B**).

Four patients in the cohort had additional variants of uncertain significance reported. Patient 7 had compound heterozygous pathogenic variants in *ACADM* associated with medium chain fatty acid dehydrogenase deficiency; NM_000016.5(ACADM):c.799G>A,p.(Gly267Arg) and NM_000016.5(ACADM):c.985A>G,p.(Lys329Glu). Patient 3 had a *de novo* Yq11.21-qter deletion and *de novo* Ypter-p11.3 duplication for which the significance is unknown. Patient 6 had a VUS in *HCFC1* associated with methylmalonic aciduria and homocysteinemia; NM_005334.3(HCFC1):c.4418C>T.p.(Thr1407Met) and patient 10 harboured a maternally inherited 16q23.3 (81,477,800 – 81,552,781) VUS.

### DDX17 supports cortical neuron development in the mouse

The identification of several variants in *DDX17* associated with neurodevelopmental features prompted assessment of DDX17 reduction on cortical development in animal models. We first turned to *in utero* cortical electroporation (IUCE) in the mouse using two distinct shRNA plasmids targeting *Ddx17* (shDDX17 #1 or shDDX17 #2). Electroporations were performed at embryonic day (E)15.5, the developmental stage at which progenitors give rise to callosal-projecting pyramidal neurons. By P21, in control conditions, all electroporated neurons (visualized by mVenus fluorescence) reached the superficial layers of the cortex (layers II/III) (**Fig 2A**). In contrast, we could observe defects in neuronal migration upon knockdown of *Ddx17* (**Fig 2B-C**) and after quantification, a statistically significant fraction of neurons did not reach the most superficial cortical layers in conditions electroporated with shRNA plasmids (**Fig 2G**). Despite this, neuronal polarization and axon formation was not impaired. In control conditions, axons of layer II/III neurons progress through the corpus callosum to reach the contralateral hemisphere, and branch extensively on ipsilateral layer V (**Fig 2A**), as well as contralateral layers II/III (**Fig 2D**). In shRNA-electroporated animals, we observed a trend toward a decrease of axon density in the ipsilateral side (**Fig 2B-C**, quantified in **Fig 2H**), and especially a strong reduction of contralateral axon density (**Fig 2E-F**, quantified in **Fig 2I**). There was no difference in axon density in the white matter (WM), indicating that the same proportion of axons reached the contralateral hemisphere regardless of *Ddx17* expression. Our results demonstrate that DDX17 is required for cortical development in the developing mouse brain.

**FIGURE 2.**
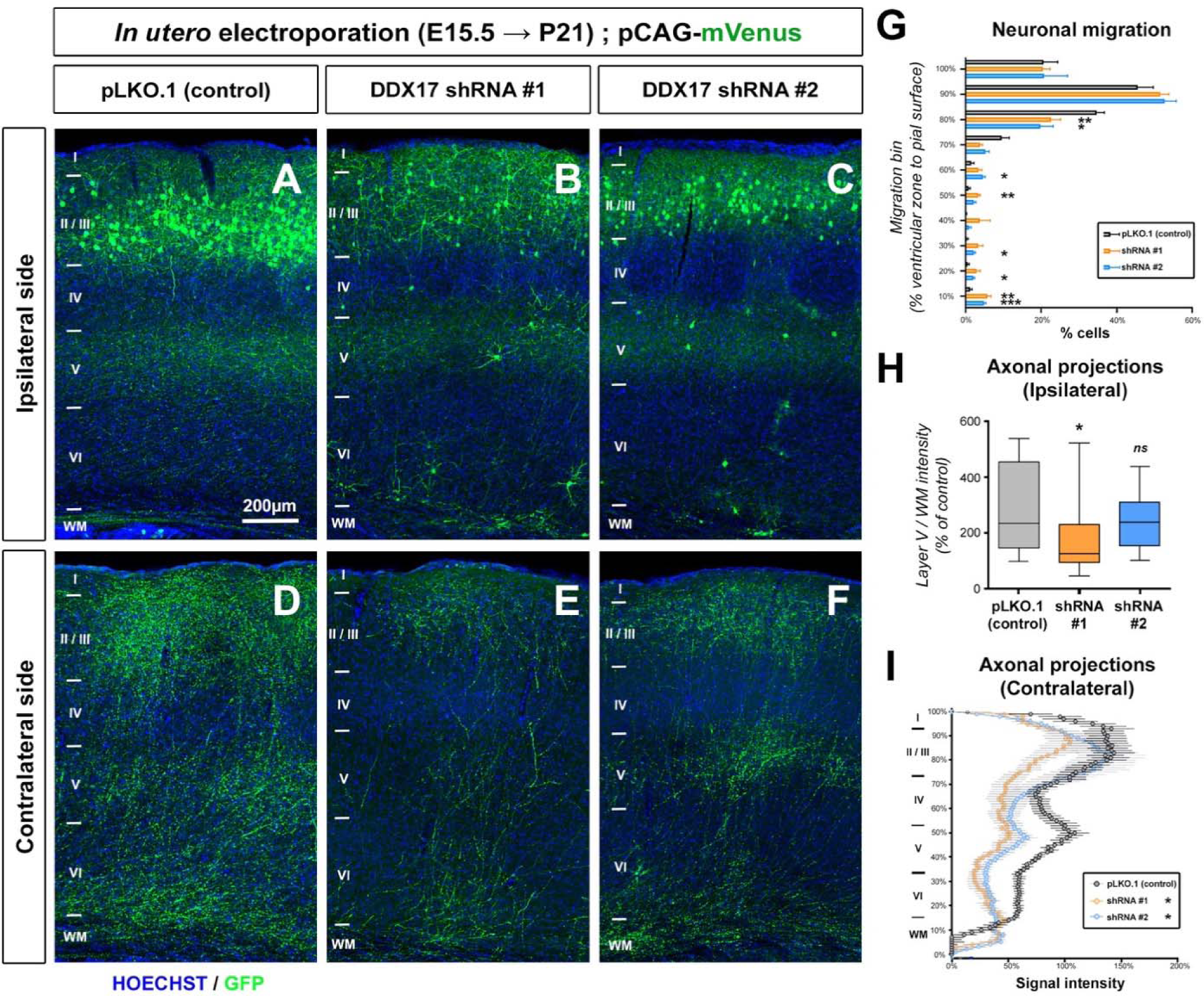
Knockdown of Ddx17 decreases cortical axon complexity in the mouse in vivo. **(A-F)** Histochemistry of the ipsilateral or contralateral side of mice at P21 following in utero electroporation with pLKO **(A,D)** or after loss of function of DDX17 (**B-C** and **E-F**) and the fluorescent protein mVenus. (**G**) Quantification of neuronal migration defects upon knockdown of DDX17. Soma position was quantified on a ventricular zone to pial surface axis. Each bin represents 10% migration. Data: average + SEM, N = 6 sections out of 3 animals (2 sections per animal). Analysis: Two-way ANOVA with multiple comparisons. *p<0.05, **p<0.01, ***p<0.001. **(H)** Quantification of normalized mVenus fluorescence in layer V of the ipsilateral cortex (min, max, median, 25^th^, and 75^th^ percentile). NpLKO=13, NshDDX17-1=18, NshDDX17-2=25. Analysis: One-way ANOVA with Dunn’s multiple comparisons. ns:p>0.05, *p<0.05. (**I**) Quantification of normalized mVenus fluorescence along a radial axis in the contralateral cortex (Average ±SEM) (H) in control condition pLKO) or after knockdown of DDX17. NpLKO=13, NshDDX17-1=18, NshDDX17-2=25. Analysis: Two-way ANOVA. *p<0.05.

Because the wiring of the brain results from sequential biological processes, defects in the early steps (e.g. neurogenesis or neuronal migration) can lead to alterations in the later biological processes such as axon development. To make sure that the axonal phenotypes observed *in vivo* do not result from abnormal neuronal migration, we subsequently turned to *in vitro* neuronal cultures, which allow quantitative assessment of axonal development at a single cell resolution. We performed *ex vivo* cortical electroporation (EVCE) at E15.5 to target neuronal progenitors in the dorsal telencephalon and cultured neurons for 5 days *in vitro* (5 DIV). In both shRNA conditions, we observed an effect on axonal development compared to the control condition (pLKO.1) (**Fig 3A-C**). The inhibition of DDX17 expression decreased axon length and reduced collateral branch formation (**Fig 3D-E**). Two independent shRNA plasmids produced markedly similar phenotypes, indicating that this phenotype is unlikely to be an off-target effect of the shRNAs. To confirm this result, we tested the consequence of overexpressing human DDX17 by EVCE. Following electroporation, we observed an increased axon length compared to the control condition, mirroring the effect of DDX17 knockdown (**Fig 3F-G**, quantified in **Fig 3H**). Interestingly, although we counted more collaterals per neuron, this increase was due to the increase in axon length and axon branching did not differ from the control condition when normalized per axon length. Overall, our results demonstrate that DDX17 is important for axon development in mouse cortical neurons.

**FIGURE 3.**
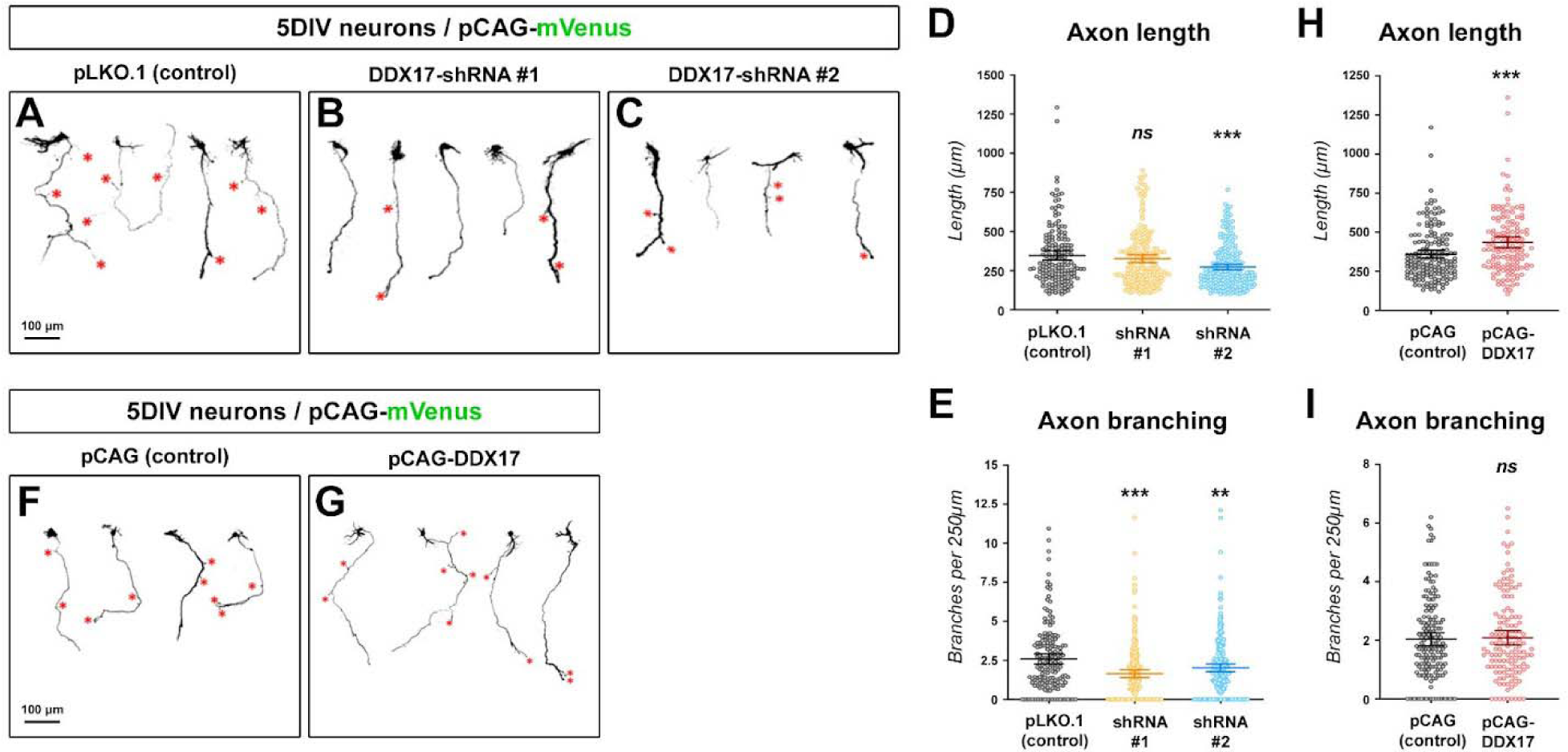
DDX17 is necessary and sufficient for axon development. **(A-C)** Representative images of mVenus expressing cortical neurons (5 DIV) in control condition (pLKO.1) or after loss of function (DDX17-shRNA #1 and DDX17-shRNA #2). Red star (*) point to collateral branches of the axon. **(D-E)** Quantification of axon length and number of collateral branches of 5 DIV neurons in the indicated conditions. Bars represent the average and 95% CI. Statistical tests: Kruskal-Wallis test with Dunn’s post-test (each condition compared to control condition). **(F-G)** Representative images of mVenus expressing cortical neurons (5 DIV) in control conditions, or upon overexpression of DDX17. Red star (*) points to branch/collateral position. **(H-I)** Quantification of axon length and number of collateral branches of 5 DIV neurons in the indicated conditions. Bars represent the average and 95% CI. Statistical tests: Kruskal-Wallis test with Dunn’s post-test. (D-E) N_(pLKO.1)_=169, N_(shDDX17 #1)_=205, N_(shDDX17 #2)_=228. (H-I) N_(pCAG)_=168, N_(pCAG-DDX17)_=134. ns: p>0.05, **: p<0.01, ***: p<0.001

### *Xenopus ddx17* crispants have reduced axon outgrowth and working memory

To test the effect of loss-of-function *ddx17* variants in an intact animal, crispant *X. tropicalis* models were used; the exon structure of the human and *Xenopus* genes are similar (**Fig 4A**) and the proteins produced have 68% amino acid identity (**Fig S2A**). The expression pattern of *ddx17* mRNA has not previously been reported in *Xenopus* and *in situ* hybridisation shows it to be expressed most highly in neural tissues including the migratory neural crest, brain, eye, and otic vesicle (see the purple staining in **Fig 4B**).

**FIGURE 4.**
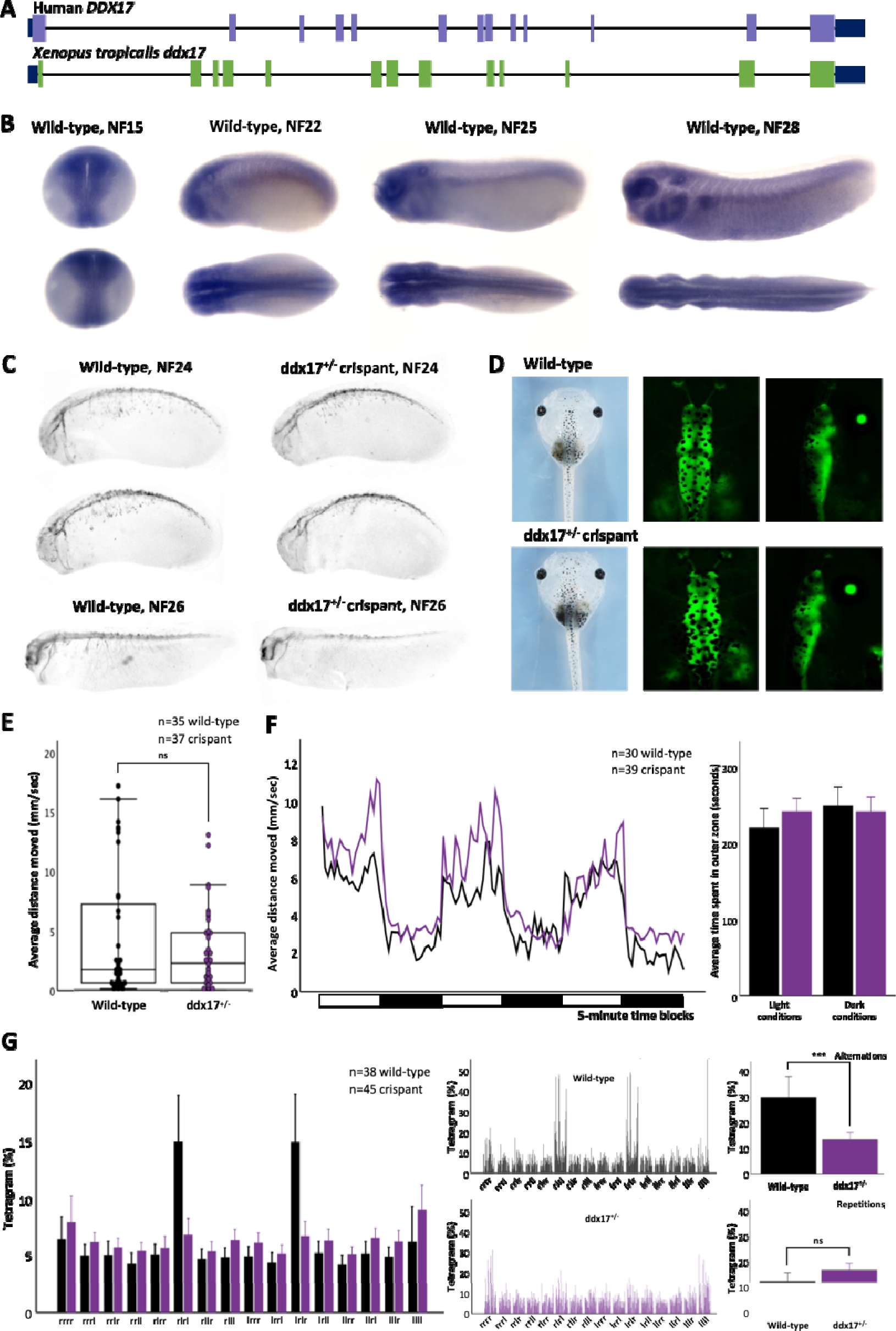
Heterozygous *ddx17 X. tropicalis* crispants appear morphologically normal but show reduced axon outgrowth and have a working memory deficit. **A.** Xenopus tropicalis and Homo sapiens have the same exon-intron structure. **B.** A developmental series of wild-type X. tropicalis were fixed and underwent in situ hybridisation with a probe specific for ddx17, the blue stain shows where this gene is expressed. The highest levels of ddx17 mRNA are in neural tissues although it is detectable more widely. **C.** Control and heterozygous crispant embryos were fixed at the stages shown and stained for neuron bodies and axons using HNK1 monoclonal antibody. The extension of axons ventrally from the neural tube is reduced in crispants at stage 24 (4/4 embryos) although growth does continue (see stage 26), scoring was blind and prior to genotyping. **D.** Brightfield microscopy showed no clear distinctions between control and crispant tadpoles across a range of stages and, when the neural tissue was labeled transgenically this too failed to reveal any gross-morphological distinctions. **E.** Tadpoles at stage NF42, similar to those shown in D, underwent automated movement analysis in a Zantiks MWP unit. In all cases the analysis was performed blind (with genotyping subsequent to measurements) the black data points represent wild-type animals with purple showing crispant data. **F.** The control and crispant animals move similar distances over time but crispants show a greater initial startle response than controls. Interestingly all tadpoles spend the majority of each 300 second block in the most proximal arena, closest to the edge of the dish (F). **G.** The main change caused by heterozygous ddx17 LOF becomes clear when working memory is tested in the free movement pattern Y-maze; the crispants have lost the alternating search pattern shown by all vertebrates.

First, a mosaic homozygous crispant knockout of *ddx17* was generated using two non-overlapping CRISPR/Cas9 gene-editing complexes designed to disrupt exon 7 (**Fig S2B-C**) and these were mated to produce F1 heterozygous and compound heterozygous tadpoles (Fig S1A). The introduction of indels into the *ddx17* locus was tested by Sanger sequencing of the target region and demonstrated strong penetrance of indels in the founder animals (**Fig S1B**), producing a *ddx17* mosaic knockout (*Xtr.ddx17*^em1E*X*RC^). The phenotypes produced by each sgRNA were indistinguishable, showing they were not due to off target effects. The heterozygous offspring produced by outcrossing mosaic founders were first identified using the T7E1 mismatch detection assay (**Fig S1C**), with the genotype confirmed by Sanger sequencing (**Fig S1D), revealing** frequently occurring deletions of 5bp, 22bp, 131bp and 136bp (**Fig S1E**), resulting in a frameshift leading to protein truncation within exon 7 (*Xtr.ddx17*^em2E*X*RC^). Inbreeding of the mosaic founders produced homozygous or compound heterozygous tadpoles as shown by Sanger sequencing (Fig S1F) (*Xtr.ddx17*^em3E*X*RC^). The homozygous tadpoles bearing larger deletions (131bp or 136bp) in one allele (**Fig S1G**), were found at later stages to have a series of further, progressive deletions as demonstrated by polyacrylamide gel electrophoresis (**Fig S1H**).

Phenotypically, more than half of the founder crispant embryos showed evidence of gastrulation defects (**Fig S3A**). The remaining animals showed no gross morphological defects (**Fig S3B,C**). The gross craniofacial morphology of *ddx17* crispants was tested by injecting CRISPR/Cas9 complexes into one-cell of a dividing two-cell embryo. This results in the effects of the protein truncation being concentrated on one side of the embryo along the left-right axis, with the other side acting as an internal control; no altered morphology was observed in the crispants (**Fig S3C**). A decrease in head size was observed prior to free-feeding stages (average 5.93 mm^2^ for control tadpoles and 4.89 mm^2^ for crispant tadpoles (NF41, n = 16), t(30)=3.85, p<0.001). This significant decrease in head size was no longer apparent at later stages of development (average 15.9 mm^2^ for control tadpoles and 16.5 mm^2^ for crispant tadpoles (NF48, n = 16), t(30=-5.05, p=0.617, **Fig S3D**).

The phenotype of patients with *ddx17* variants include neurodevelopmental deficits and these can now be modeled in *X. tropicalis*. Here, in addition to the Free-movement Pattern (FMP) Y-maze that is already validated in *X. tropicalis,*^20^ another assay has been adapted to assess working memory in *Xenopus*. In this assay, animals are exposed to 5-minute alternating periods of light and dark; their initial response is to move (startle) as the lights go off, but they quickly learn that the dark period does not signal danger and reduce their response to it. This attenuated response to the changing environmental conditions demonstrates a second measure of short-term working memory in the tadpoles. Importantly this behavior can be probed by pharmacological agents which disrupt working memory and attenuate anxiolytic responses. Following treatment with the NMDA receptor antagonist MK-801, tadpoles can be observed to startle repeatedly in response to the light-dark transition period. The startle response is also abolished following administration of 5 μM diazepam (**Fig S4A**). Once established using pharmacological agents, these assays were applied to test the neurodevelopmental phenotypes of the *ddx17* crispants.

Comparing the movement of control tadpoles and F_0_ *ddx17* crispant animals at NF48 showed that crispant animals move less over the 10-minute trial period (average 4.0 mm/sec for control tadpoles and 2.7 mm/sec for crispant tadpoles (n = 48, t(82)=-2.6, p=0.012), **Fig S3F**). A significant reduction in locomotion was additionally observed in a second assay investigating the tadpoles’ response to 5-minute light-dark transition periods (average 3.9 mm/sec for control tadpoles and 2.3 mm/sec for crispant tadpoles (n = 24, t(46)=3.4, p=0.001), **Fig S3G**). Unusually, these mosaic crispant tadpoles were not observed to startle at these light-dark transitions. In the FMP Y-maze assay that tests working memory, founder crispant tadpoles performed fewer alternations in their search patterns than controls (average alternations 22.8% for control tadpoles and 19.5% for crispant tadpoles (n = 20, F(1, 33) = 0.366, p = 0.550)) and more repetitions (average repetitions 9% for control tadpoles and 12.8% for crispant tadpoles (n = 20, F(1, 33) = 3.272, p = 0.080)) (**Fig S3H**).

Since there was clear evidence of movement and neural defects in the F_0_ crispants, together with reduced axon length observed in electroporated mouse brains deficient in DDX17, neuron outgrowth was examined in *Xenopus* using an anti-HNK-1 antibody. Embryos injected with control (*tyr*)^21^ and *ddx17* gene-editing complexes restricted to one side of the embryo reveal reduced axon outgrowth in *ddx17* crispants on the ipsilateral side (compare the injected and uninjected lateral views in **Fig S4E**). Similarly, in the heterozygous *ddx17* model axon outgrowth is visibly reduced compared with controls at NF stage 24 with evidence of continued axon outgrowth at later stages (NF26 onwards (**Fig 4C & Fig S4B**)).

In comparison to the founder animals, F_1_ crispant tadpoles bearing non-mosaic heterozygous or compound heterozygous/homozygous indels in *ddx17* show no gross morphological or developmental abnormalities including in gastrulation (**Figs 4D, S4C & S5A**), hatching rate and head size (**Fig S5B**). Gross structural differences between control and crispants were not seen in the forebrain, midbrain or hindbrain regions of heterozygous *ddx17* crispant animals, even when bred in a [Xtr.Tg(tubb2b:GFP)Amaya] RRID: EXRC_3001 background (**Fig S4C**) to make differences more obvious. Post-hatching, all animals moved away from tactile stimuli (to the head) and at later stages adopted a normal, head down filter-feeding posture with the ability to navigate freely within their respective environments. Further, no abnormalities in locomotive behavior consistent with descriptors of seizure activity in tadpoles were observed.^22,23^ The locomotive activity of wild-type tadpoles, heterozygous or homozygous, non-mosaic crispants in *ddx17* was indistinguishable at NF48 when tracked across a 10- minute trial period (**Fig 4E & S5C**). Similarly, comparative average locomotive activity in the light-dark assay was not significantly different from controls in either non-mosaic crispant group, with all tadpoles noted to spend the majority of each of the 5-minute blocks at the edge of the dish.

Unlike wild-type tadpoles however, both non-mosaic crispant tadpole models were noted to respond with increased locomotion to multiple dark-light transition periods, although startling was more pronounced in the homozygous/compound heterozygous model (**Fig 4F & S5D**). Since this indicated reduced memory, we used a well characterized working memory assay, the 1 hour free-movement pattern Y-maze to compare wild-type and homozygous or heterozygous *ddx17* crispant tadpoles. Although not significant, homozygous *ddx17* tadpoles performed fewer alternations (average alternations 22.6% for control tadpoles and 15.2% for homozygous *ddx17* tadpoles (n = 49, F(1, 95) = 3.761, p = 0.055)) but significantly more repetitions (n = 49, F(1, 95) = 10.983, p = 0.001) than controls (**Fig S5E**). This increase in repetitions taken together with the unattenuated startle response suggests increased levels of anxiety in the homozygous *ddx17* tadpole group.^24^ In comparison, the heterozygous *ddx17* model demonstrated significantly fewer alternations (F(1, 80) = 14.25, p < 0.001) without a significant increase in repetitions (F(1, 80) = 3.29, p = 0.074) when compared to wild-type tadpoles (**Fig 4G**). When considered alongside the response observed in the light-dark assay, this shows a clear deficit in the short-term working memory.

Overall, the crispant *ddx17* models show very similar phenotypes, with evidence of reduced axon outgrowth and working memory consistent between them. The mosaic founder animals however move less than the wild-type tadpoles unlike the other crispant models and have a transiently reduced head size.

### RNA-seq analysis

Finally, to gain insight into the possible functions and target genes of DDX17 in a human cellular context, we carried out a transcriptomic analysis of neuroblastoma SH-SY5Y cells in which we knocked-down the expression of the *DDX17* gene, using a mixture of 2 different siRNAs (**Fig 5A**). We identified 350 genes that were differentially expressed in *DDX17*-KD cells compared to control cells (**Fig 5B** and **Supplementary File C**). The functions of this set of genes were significantly associated to developmental processes, in particular the development and functions of the nervous system (**Fig 5C, S6,** and **Supplementary File A**). For instance, the expression of several development-associated transcription factors (*MSX2*, *TBX3*, *GATA3*, *FOSL1*, *NEUROG2*, *SMAD6* and *SMAD9*, *SOX13*, *DRGX*, *RARB*, *MYCN…*) was deregulated upon *DDX17* KD. Of note, looking at the molecular functions associated with those genes also revealed the presence of a significant number of *trans*-membrane receptors (31 genes) and receptor ligands (15 genes) (**Supplementary File C**), including several receptors/ligands associated with axon guidance (*DCC*, *EFNB2*, *PLXNA2* and *PLXNA4*, *SEMA6A* and *SEMA6D*, *RET*, *ROBO2*, *UNC5D*…).

**FIGURE 5.**
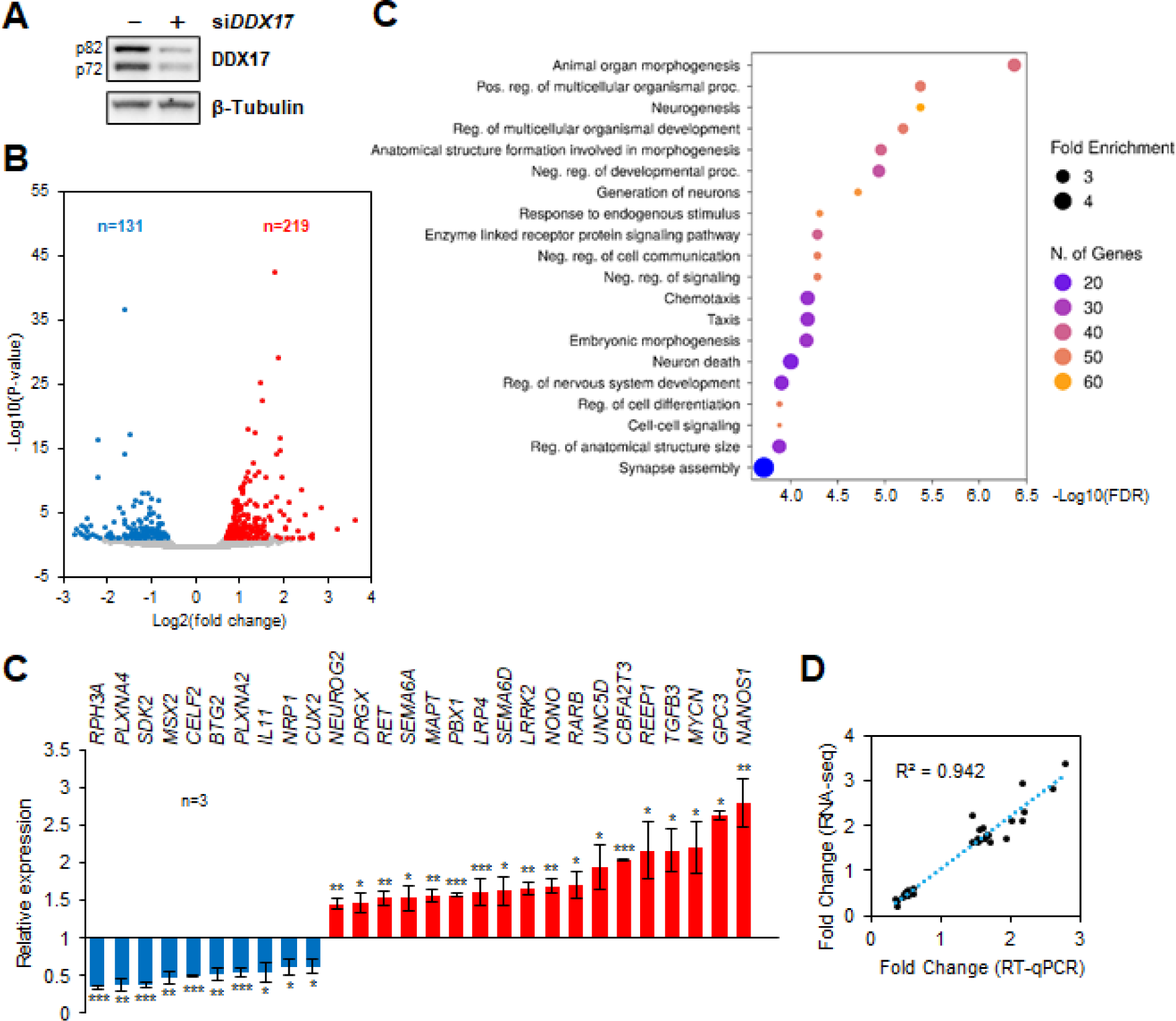
DDX17 controls the expression of genes involved in nervous system development. **A.** Western-blot showing the siRNA-mediated depletion of DDX17 protein in SH-SY5Y cells. B. Volcano plot showing the genes that are impacted by DDX17 KD in SH-SY5Y cells, as predicted from the RNA-seq analysis. Significantly altered genes (downregulated in blue and upregulated in red) were identified as described in the Methods section. **C.** Gene ontology analysis using ShinyGO for the genes impacted by DDX17 KD. Only the top 20 of the GO enriched biological processes are shown (see Supplementary File A for the full list of enriched terms). **D.** Validation of the effect of DDX17 knockdown on the steady-state expression of a selection of genes. RT-qPCR data were first normalized to GAPDH mRNA level in each condition, and the normalized mRNA level of each gene in the DDX17 knockdown condition was then normalized to the control condition, set to 1. Data are expressed as the mean value ± S.E.M. of independent experiments (n = 3). Unpaired Student’s t-test (*P-val < 0.05; **P-val < 0.01; ***P-val < 0.001). **E.** Correlation between the measured fold change of expression (x-axis) and the corresponding predicted fold change value (y-axis) for the 28 genes shown in panel E.

We then analyzed separately the subsets of genes of which steady-state expression was negatively (131 genes) or positively (219 genes) altered by *DDX17* KD (**Supplementary files A & C**). This analysis showed again a significant occurrence of GO terms associated with neurogenesis in both groups of genes, but it also underlined a link between downregulated genes and body morphogenesis, while the group of upregulated genes was associated more specifically to cell-signaling pathways (**Fig S7**). To validate our computational analysis, we selected 28 genes from the two subgroups of mis-regulated genes and measured their mRNA level by RT-qPCR assays in mock-depleted and DDX17-depleted SH- SY5Y cells. Consistently, the expression of each tested gene was altered as predicted from the RNA-seq, with a strong combined correlation score (R^2^=0.942) (**Figs 5D** and **5E**).

Collectively, our results indicate that the *DDX17* gene is involved in several processes during the development of vertebrates, in particular in the development of the nervous system, most likely because they control the expression of subsets of genes with key functions in the neural system.

## Discussion

To the best of our knowledge, this is the first study to describe *de novo* heterozygous *DDX17* variants associated with features describing a novel neurodevelopmental disorder. Using mouse and frog animal models, we provide strong evidence that DDX17 plays important roles in the developing nervous system, in mouse and frog models. More specifically, *Ddx17* knockdown impaired neuronal migration and axon development in the brain of newborn mice and reduced axon outgrowth and branching in primary cortical neurons *in vitro*. In agreement with these results, crispant tadpole *ddx17* models, including a heterozygous F_1_ model, also presented a reduced axon outgrowth phenotype. Crispant tadpoles also had clear functional neural defects. Since the region and developmental state of the central nervous system in the mice and tadpoles in which this effect has been noted are distinct, it suggests that *DDX17* has an important role in widely distributed neurodevelopmental processes. The conservation of function across evolutionarily distant species such as mice and frogs strongly support that the role of *DDX17* is conserved in humans too. These *in vivo* results are therefore consistent with the hypothesis that heterozygous loss-of-function *DDX17* variants identified in patients induce a significant alteration of the function of the protein during neuronal development, resulting in the observed phenotype in our cohort.

The phenotype associated with *de novo* variants in *DDX17* is consistent with a neurodevelopmental disorder, typified by mild-moderate intellectual disability, delayed speech and language development and global developmental delay. Sixty-four percent (7/11) of the cohort have dysmorphic facial features, although for many this is subtle. Overlapping dysmorphology between patients includes: synophrys; upslanting palpebral fissures; depressed nasal bridge; posteriorly rotated ears; high arched eyebrows; epicanthus; telecanthus; and strabismus. Some patients have gross and fine motor delay, generalized hypotonia, sterotypy, and evidence of autism spectrum disorder.

There were no substantial differences in phenotype severity between patients harboring missense variants versus loss-of-function variants, suggesting DDX17 haploinsufficiency causes the observed phenotype. Since all missense variants but one fall within the helicase domain (**Fig 1**), it suggests that they impact the structure and/or activity of this domain in a way that deeply alters the overall function of DDX17, similarly to loss-of-function variants. Our preliminary molecular modelling analyses did not reveal any significant modification of the DDX17 structure in which patient missense mutations were introduced (data not shown). However, our analysis was based on the only known 3D structure of human DDX17, which is limited to the helicase core domain^25^. Recently it has been shown that the two disordered and flexible flanking domains also strongly affect the helicase activity of Dbp2, the yeast DDX17 ortholog.^26^ It is thus currently impossible to accurately predict the impact of variants on DDX17 function, without taking into account the interactions between the structured and unstructured regions of the protein.

We, and others^6,7^ have shown previously a role of DDX17 in the retinoic acid induced differentiation of neuroblastoma SH-SY5Y and pluripotent embryonal NTERA2 cells, respectively. However, this effect was most evident when DDX17 knockdown was combined with the concomitant depletion of its paralog DDX5. We now demonstrate that the downregulation of DDX17 alone is sufficient to alter neuronal development, both *in vivo* and *in vitro*. Both DDX17 and DDX5 have largely redundant functions, which probably explains why their joint depletion has such a strong effect compared to single protein depletion.

Interestingly, two distinct shRNAs targeting DDX17 alter neuronal migration in the mouse cortex. Although shRNA-based strategies are prone to off-target disruption of neuronal migration in the mouse cortex^27^, the migration phenotype is compatible with our previous observation that DDX17 controls the activity of the Repressor Element 1-silencing transcription factor (REST) complex during neurogenesis^6^ and that the REST/CoREST complex regulates neuronal migration.^28^ Future studies using genetic knockout models will demonstrate the specificity of the migration phenotype. Furthermore, we report that DDX17 plays a role in axon morphogenesis that is independent of its function in neuronal migration.

*Xenopus* frogs have been used as pioneer model organisms since the mid-twentieth century, mainly in discovery research^29^. Gene editing was found to be exceptionally effective in them and their application as tools for studying disease has increased.^30^ *X. tropicalis* are diploid tetrapods with very few gene duplications. Their genome structure has high levels of synteny with humans^31^ and the initial determination that 80% of human disease genes have orthologues in this species is now thought to be an underestimate.^32^ We and others have shown them to be highly suited to testing the links between a variant of uncertain significance and human disease phenotypes.^20,33^ This can often be achieved without breeding the animals due to the efficiency of CRISPR/Cas resulting in very low levels of mosaicism in founders. Hence, they represent a rapid and cost-effective assay for gene-disease associations, filling an important gap between the mouse and zebrafish models. Here we have used mosaic founders, and both heterozygous and compound heterozygous/homozygous F_1_ non-mosaic models to test the effect of a truncation in ddx17. Here, we have expanded the use of *Xenopus* in disease modelling; we applied a behavioral assay to tadpoles (the light dark-transition assay) that has been used previously in other models. We then confirmed the measurements of both working memory and anxiety with small molecule inhibitors. Additionally, we directly compared mosaic homozygous F_0_ crispants with non-mosaic F_1_ homozygous and F_1_ heterozygous animals. There were stronger phenotypic effects in mosaic founder animals. This is a known phenomenon, which may be associated with a failure to activate compensatory mechanisms in mosaic animals, including crispants in another aquatic model, zebrafish (reviewed by Rouf *et al*.^34^). This suggests that *Xenopus* behave like zebrafish in this respect.

Our data offer some limited insights into the mechanism whereby DDX17 variants affecting its function relate to the disease phenotype. Since DDX17 is known to regulate gene expression at multiple levels, the different pathological features associated with *DDX17* mutations likely result from the altered expression of some of its target genes and transcripts. Indeed, our transcriptomic analysis showed that 350 genes may be impacted, a large proportion of which are important for development and morphogenesis, and most particularly for neurogenesis. This includes several key transcription factors (*NEUROG2*, *RARB*, *MYCN…*), the deregulation of which could have direct and indirect effects on many other genes in the course of embryonic development. Furthermore, the DDX17-dependent regulation of several genes coding for trans-membrane receptors and ligands associated with axon guidance is also of particular significance, considering the altered axonal development observed upon DDX17 knockdown in mice and tadpoles, and the neurological phenotype observed in patients. Whilst further work is needed, the goal of this study is to establish DDX17 as a novel neurodevelopmental disease gene and enable identification of more patients to further elucidate the genotype-phenotype relationship.

## Conclusion

We have identified 11 patients with neurodevelopmental phenotypes harbouring monoallelic *de novo* variants in *DDX17*. Functional experiments (*in vitro* and *in vivo*) show that *DDX17* is important in neurodevelopmental processes, in keeping with the observed human phenotype. *Ddx17* knockdown of newborn mice showed impaired axon outgrowth, and reduced axon outgrowth and branching was observed in primary cortical neurons *in vitro*. The axon outgrowth phenotype was replicated in crispant *ddx17* tadpoles, including in a heterozygous (F_1_) model. Crispant tadpoles had clear functional neural defects and showed an impaired neurobehavioral phenotype. Transcriptomic analysis further supports the role of *DDX17* in neurodevelopmental processes, particularly neurogenesis. These results strongly support that monoallelic loss-of-function variants in *DDX17* cause a neurodevelopmental phenotype.

## Data Availability

Transcriptomic data were deposited in the Gene Expression Omnibus (GEO) database under the record GSE223072. The following secure token has been created to allow review of record while it remains in private status: wrwfgmgibzedhwx. The embargo will be released upon acceptance of the manuscript. The published article includes all remaining data generated or analyzed during this study.

## Acknowledgements

Claudia Ciaccio and Stefano D’Arrigo are members of the ITHACA-ERN. Mouse experiments were performed with support from the Service Commun des Animaleries de Rockefeller (SCAR) from the University of Lyon.

